# Regressing SARS-CoV-2 sewage measurements onto COVID-19 burden in the population: a proof-of-concept for quantitative environmental surveillance

**DOI:** 10.1101/2020.04.26.20073569

**Authors:** Itay Bar-Or, Karin Yaniv, Marilou Shagan, Eden Ozer, Oran Erster, Ella Mendelson, Batya Mannasse, Rachel Shirazi, Esti Kramarsky-Winter, Oded Nir, Hala Abu-Ali, Zeev Ronen, Ehud Rinott, Yair E. Lewis, Eran Friedler, Eden Bitkover, Yossi Paitan, Yakir Berchenko, Ariel Kushmaro

## Abstract

SARS-CoV-2 is an RNA virus, a member of the coronavirus family of respiratory viruses that includes SARS-CoV-1 and MERS. COVID-19, the clinical syndrome caused by SARSCoV-2, has evolved into a global pandemic with more than 2,900,000 people infected. It has had an acute and dramatic impact on health care systems, economies, and societies of affected countries within these few months. Widespread testing and tracing efforts are employed in many countries in order to contain and mitigate this pandemic. Recent data has indicated that fecal shedding of SARS-CoV-2 is common, and that the virus can be detected in wastewater. This indicates that wastewater monitoring is a potentially efficient tool for epidemiological surveillance of SARS-CoV-2 infection in large populations at relevant scales. Collecting raw sewage data, representing specific districts, and crosslinking this data with the number of infected people from each location, will enable us to derive and provide quantitative surveillance tools. In particular, this will provide important means to (i) estimate the extent of outbreaks and their spatial distributions, based primarily on in-sewer measurements (ii) manage the early-warning system quantitatively and efficiently (and similarly, verify disease elimination). Here we report the development of a virus concentration method using PEG or alum, providing an important a tool for detection of SARS-CoV-2 RNA in sewage and relating it to the local populations and geographic information. This will provide a proof of concept for the use of sewage associated virus data as a reliable epidemiological tool.

## Introduction

Waterborne pathogens, including viruses, bacteria and protozoa can be shed into the urban water cycle *via* sewers, (Gormley et al. 2017 and 2020) urban runoff, agricultural runoff and wastewater discharges (Arnone and Walling, 2007; La Rosa et al., 2012). Indeed, it has been reported that there are high concentrations of virus particles in wastewater treatment plants (WWTP), varying from 10^8^ to 10^10^ viral particles per milliliter (Otawa et al., 2007). Coronavirus SARS-CoV-2 is a novel RNA virus belonging to a group of viruses that includes amongst others SARS and MERS. SARS-CoV-2 is one of more than 37 coronaviruses in the *Coronaviridae* family, within the order *Nidovirales*, and it is currently causing a major pandemic with over 2,900,000 people infected globally. It causes COVID-19, a disease that has daunting effect on health care systems, economies, and societies of affected countries. As a member of the *Coronaviridae*, which includes viruses known to cause respiratory and/or intestinal infections, SARS-CoV-2 spreads primarily via micro droplets, reflecting its survivorship in humid environments (Chin et al. 2020). Recent reports have detailed SARSCoV-2 shedding in human stool (Gao et al. 2020; Hindson 2020; Xu et al. 2020). Interestingly it has been demonstrated that a similar corona virus, SARS-CoV-1 can survive in sewage for 14 days at 4°C, and for 2 days at 20°C, and its RNA can be detected for 8 days, even though the virus was inactive (Gundy et al. 2009; Wang et al. 2005). In recent studies, the SARS-CoV-2 virus was reported to be present in wastewater in treatment facilities (Hindson et al. 2020; Medema et al. 2020; Naddeo and Liu 2020; Wurtzer et al. 2020; Wu et al. 2020). Despite this, we are still lacking sufficient studies regarding the fate of SARSCoV-2 throughout the different stages of wastewater collection and treatment.

Presence and prevalence of SARS-CoV-2 in wastewater provides a valuable epidemiological data source (Lodder & de Roda Husman, 2020). Wastewater-based epidemiology (WBE) is a new discipline concerned with mining chemical and biological information from municipal wastewater. WBE has been applied for populations around the globe to measure chemicals consumption and exposure patterns (Choi et al. 2019). It was proven to be useful for preclinical identification (i.e., before the population exhibited symptoms) of Aichi virus (Lodder & de Roda Husman, 2020), for monitoring antibiotic resistance on a global scale (Lodder & de Roda Husman 2020), for quantitative polio surveillance (Berchenko et al. 2017), and also as fecal indicators (Gu et al. 2018, Saeidi et al. 2018). In particular, in our previous work, (Berchenko et al. 2017) we obtained valuable epidemiological information regarding polio by analyzing two unique data sets collected during a “natural experiment” provided by the 2013 polio outbreak in Israel (wastewater data from different locations, and records of supplemental immunization with the live vaccine). The parametric characterization of the linear dose-dependent relationship between the number of poliovirus shedders and the amount of poliovirus in sewage yielded a powerful tool for quantitative environmental surveillance (Berchenko et al. 2017). Here we report a similar study aimed at developing similar quantitative tools for SARS-CoV-2 in wastewater. These results will enable early identification and spatial-based monitoring of future outbreaks, and could be used to confirm virus elimination or to validate the need for more containment efforts.

## Material and methods

### Sampling

Samples were taken from wastewater treatment plants in different locations in Israel (see Table 2) as well as samples of raw sewage from different districts from the Tel Aviv metropolis. Sampling equipment was sanitized and properly sterilized (cooler, sampling bottles, biohazard bags, etc.). In addition, we used automatic samplers at targeted hot-spot areas for 24 hours. Around 200 ml were collected every 30 min for the 24 hours. Samples from the automatic sampler (6-10 L) were transported immediately to the lab where samples were poured to 2L of clean plastic bottles. Fresh 1 ml of raw sewage was taken directly into lysis buffer for RNA extraction. The rest of the sample was stored at −20°C or −80°C for virus concentration and RNA extraction stages.

### Sample concentration and analysis

Viral particles from approximately 0.25-1 liter of sewage/wastewater /effluent samples were concentrated using first centrifugation to remove sediment and large particles. Secondary concentration of the supernatant was performed using polyethylene glycol (PEG) or alum (20mg/l) precipitation, followed by additional centrifugation. The mixture was incubated at 4°C with 100-rpm agitation for about 12h, then centrifuged at 14,000 g for 45 min at 4°C to pellet the virus particles. Virus particles were then resuspended in phosphate buffered saline (PBS). The aqueous phase (containing virus particles) was collected and filtered through a 0.22µm filter. Ultra-15 centrifugal tubes with a molecular weight cutoff of 30 kDa (Amicon) were used to further concentrate the sample to a final volume of 1 ml. Samples were stored at −20/−80°C until further analysis. After primary and secondary concentrations, viral RNA was extracted from the samples using viral RNA extraction kit (RNeasy mini kit- QIAGEN and EasyMAG -bioMerieux, France) and then stored at - 80°C.

### Identification and quantification of Corona virus

The extracted viral nucleic acids were reverse transcribed and qPCR for the cDNA was performed using mixture containing Fast Start Universal Probe Master, forward and reverse primers including TaqMan probe. Duplicate qPCR amplification was performed in a Step One Plus real-time PCR system. Both positive-control and negative-control assays were performed for quality control. Serial dilutions of plasmid DNA containing the SARS-CoV-2 E gene used to generate the standard curve.

## Results and discussion

This study enabled us to establish novel virus concentration methods (Tables 1 & 2). These methods were validated using sewage samples from the Dan Panorama Hotel in Tel Aviv, which currently functions as a COVID-19 isolation facility. Results show that the most efficient concentration methods were achieved using PEG and Alum reaching positive values with qPCR Ct of 33 and 33.6 for PEG and Alum, respectively (Table 1). We further established a proof-of-concept for our ability to detect SARS-CoV-2 RNA from raw sewage samples from different location in Israel (Table 3). We found traces of the virus in sewage originating from the Sorek wastewater treatment plant (Ct 32.9) as well as from Bnei Brak city sewage sampling points (Ct 33-37). The concentration of the virus RNA (as Ct) in the Bnei Brak sewage (the sampling points do not cover all the sewage distribution system in the city) correlated with the general number of COVID-19 positive individuals in the city (see Figure 1). Furthermore, the change observed in the Jerusalem sampling points from the end of March to 21 to April may demonstrate the dynamics of COVID-19 outbreak in the community. Interestingly the Beer Sheva, and Haifa samples were negative (>Ct 40) for SARS-CoV-2, possibly related to the low proportion of infected people in these cities (Table 3).

**Table 1.** SARS-CoV-2 virus concentration methods from sewage

| Sample | Method (reference) | Ct value |  | Interpretation |
| --- | --- | --- | --- | --- |
| | | FAM (SARS-CoV-2) | VIC (Internal reference β-actin) | |
| Positive Control | BGI kit (Real Time Fluorescence RT-PCR) | 27.4 | 27.3 | Positive |
| TLV1 pellet | Kazama et al. 2016 | 32.8 | 31.0 | Positive |
| TLV1 supernatant |  | 33.1 | 31.9 | Positive |
| TLV2 | Mathis et al. 2017 | Undetermined | 34.5 | Negative |
| TLV3 | Gu et al, 2018 | Undetermined | 36.4 | Negative |
| TLV4 pellet | 20mg alum per liter | 33.6 | 31.6 | Positive |
| TLV4 supernatant |  | 35.4 | 35.7 | Negative |

**Table 2:**
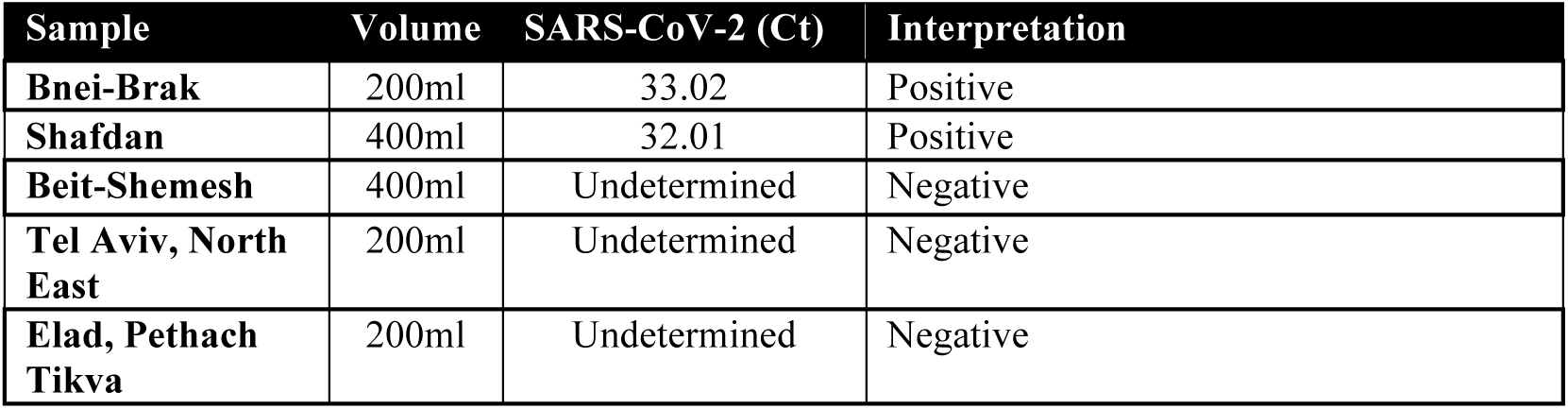
SARS-CoV-2 detection using Alum concentration method and spin column RNA mini kit

**Table 3.**
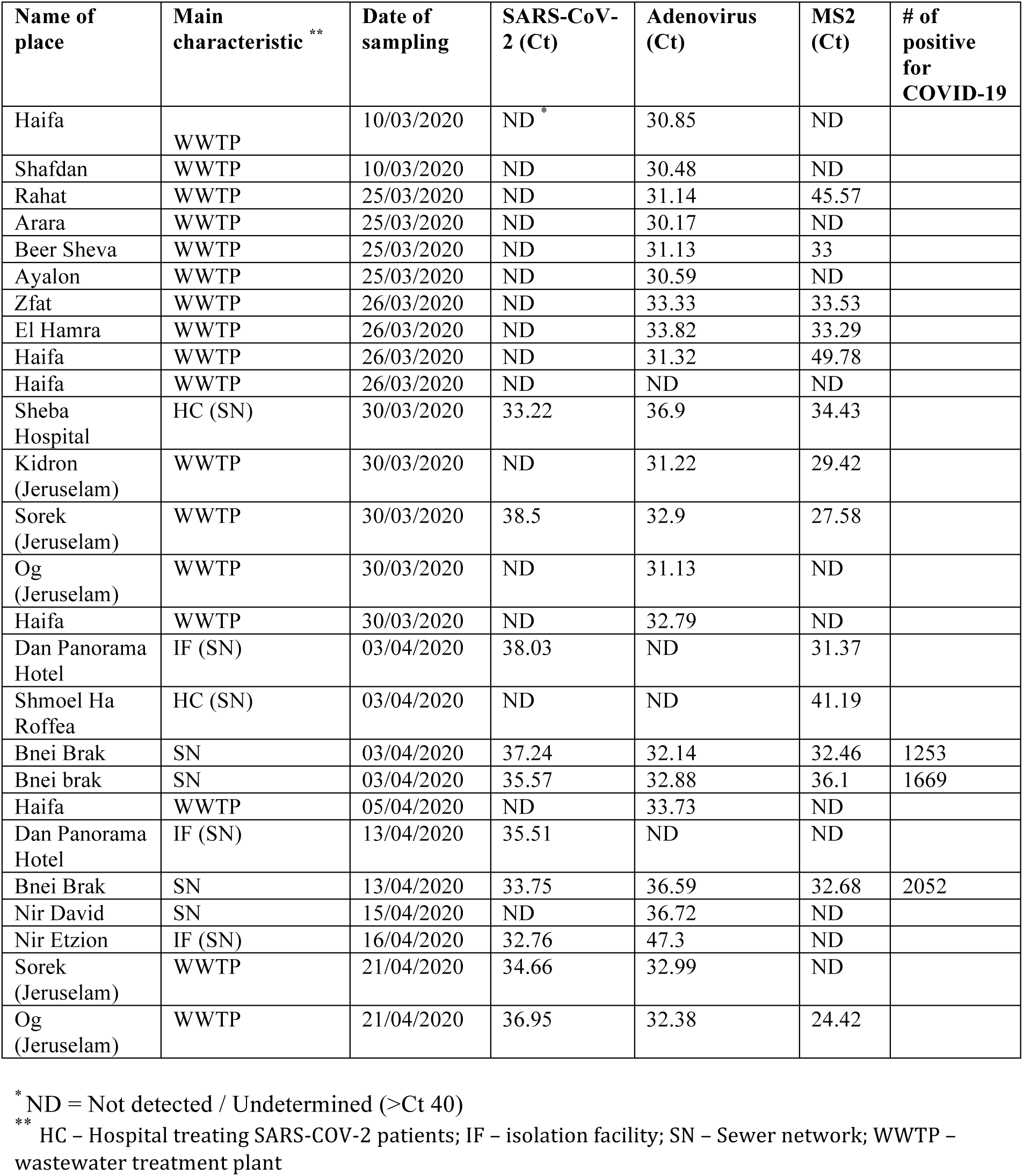
Proof-of-concept for detecting SARS-CoV-2 from raw sewage in different localities in Israel

| Name of place | Main characteristic ** | Date of sampling | SARS-CoV-2 (Ct) | Adenovirus (Ct) | MS2 (Ct) | # of positive for COVID-19 |
| --- | --- | --- | --- | --- | --- | --- |
| Haifa | WWTP | 10/03/2020 | ND * | 30.85 | ND |  |
| Shafdan | WWTP | 10/03/2020 | ND | 30.48 | ND |  |
| Rahat | WWTP | 25/03/2020 | ND | 31.14 | 45.57 |  |
| Arara | WWTP | 25/03/2020 | ND | 30.17 | ND |  |
| Beer Sheva | WWTP | 25/03/2020 | ND | 31.13 | 33 |  |
| Ayalon | WWTP | 25/03/2020 | ND | 30.59 | ND |  |
| Zfat | WWTP | 26/03/2020 | ND | 33.33 | 33.53 |  |
| El Hamra | WWTP | 26/03/2020 | ND | 33.82 | 33.29 |  |
| Haifa | WWTP | 26/03/2020 | ND | 31.32 | 49.78 |  |
| Haifa | WWTP | 26/03/2020 | ND | ND | ND |  |
| Sheba Hospital | HC (SN) | 30/03/2020 | 33.22 | 36.9 | 34.43 |  |
| Kidron (Jerusalem) | WWTP | 30/03/2020 | ND | 31.22 | 29.42 |  |
| Sorek (Jerusalem) | WWTP | 30/03/2020 | 38.5 | 32.9 | 27.58 |  |
| Og (Jerusalem) | WWTP | 30/03/2020 | ND | 31.13 | ND |  |
| Haifa | WWTP | 30/03/2020 | ND | 32.79 | ND |  |
| Dan Panorama Hotel | IF (SN) | 03/04/2020 | 38.03 | ND | 31.37 |  |
| Shmoel Ha Roffea | HC (SN) | 03/04/2020 | ND | ND | 41.19 |  |
| Bnei Brak | SN | 03/04/2020 | 37.24 | 32.14 | 32.46 | 1253 |
| Bnei brak | SN | 03/04/2020 | 35.57 | 32.88 | 36.1 | 1669 |
| Haifa | WWTP | 05/04/2020 | ND | 33.73 | ND |  |
| Dan Panorama Hotel | IF (SN) | 13/04/2020 | 35.51 | ND | ND |  |
| Bnei Brak | SN | 13/04/2020 | 33.75 | 36.59 | 32.68 | 2052 |
| Nir David | SN | 15/04/2020 | ND | 36.72 | ND |  |
| Nir Etzion | IF (SN) | 16/04/2020 | 32.76 | 47.3 | ND |  |
| Sorek (Jerusalem) | WWTP | 21/04/2020 | 34.66 | 32.99 | ND |  |
| Og (Jerusalem) | WWTP | 21/04/2020 | 36.95 | 32.38 | 24.42 |  |

\* ND = Not detected / Undetermined (>Ct 40)
\*\* HC – Hospital treating SARS-COV-2 patients; IF – isolation facility; SN – Sewer network; WWTP – wastewater treatment plant

**Figure 1.**
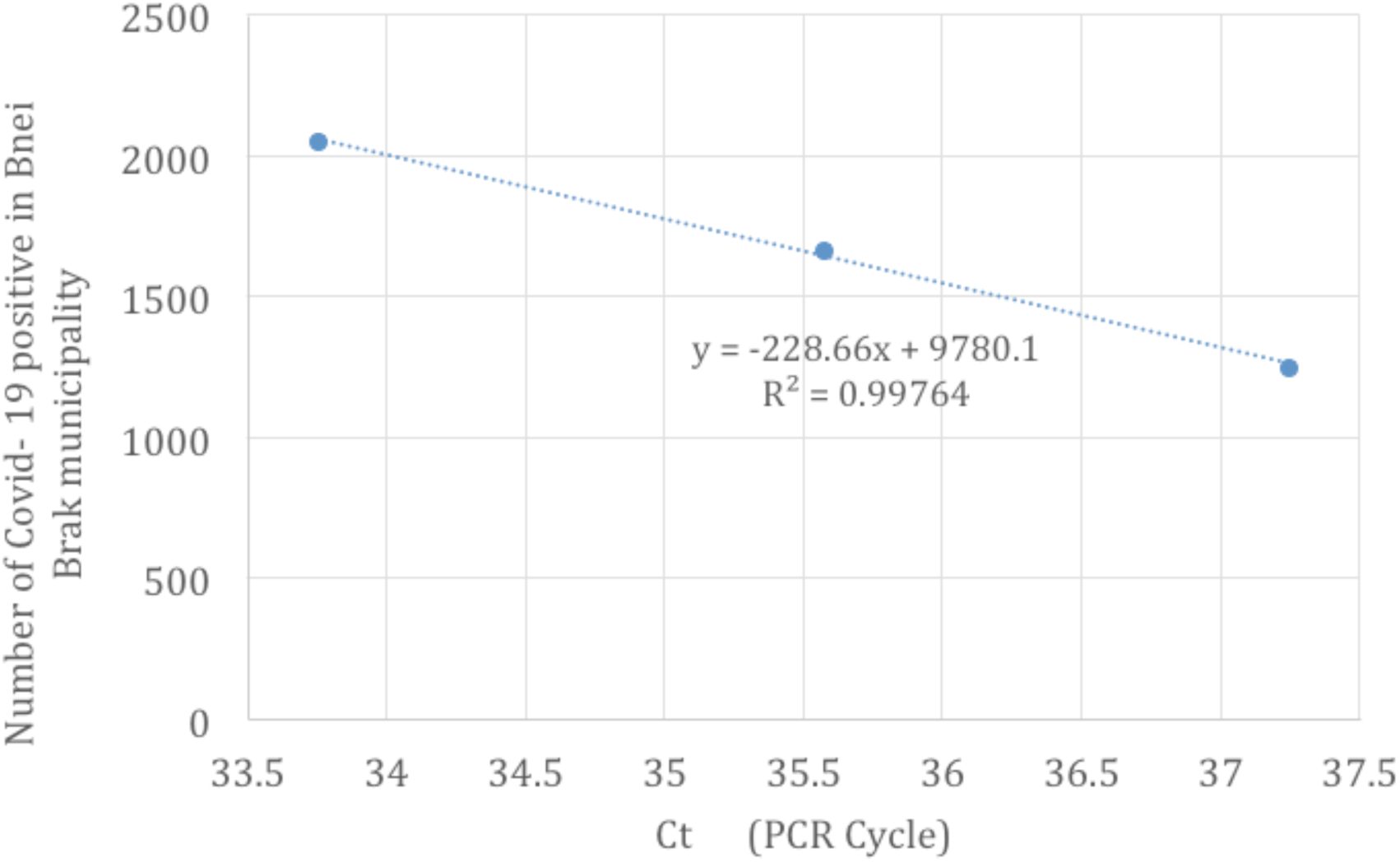
SARS-CoV-2 Ct in raw sewage (qPCR) verses number of positive diagnosed Covid-19 in Bnei Brak city.

In conclusion, we present a preliminary study demonstrating a proof-of-concept for the detection of SARS-CoV-2 RNA in sewage. We also present a linear “dose-dependent” curve as a tool for viral surveillance in environmental samples. However, we urge the readers to be cautious in their use of fig. 1 as a basis for their own data since: (i) our previous work (Berchenko et al. 2017) showed how different localities should be compared by considering daily sewage production as a measure of the local population size, and (ii) our work is still preliminary and ongoing, therefore further data is still warranted. Furthermore, we present two methods for viral isolation from wastewater by concentration with PEG and/or alum. Understanding the ecology of SARS-CoV-2 in human waste could lead to environmental applications including efficient monitoring and surveillance of SARS-CoV-2. In the future, this study may also provide tools to for sewage monitoring as an early warning alarm for SARS-CoV-2 outbreaks in the population.

## Data Availability

All data is included in the tables and figures

## Acknowledgments

We would like to thank funding from Ben Gurion University, The Corona Challenge Covid-19 (https://in.bgu.ac.il/en/corona-challenge/Pages/default.aspx) and to the wastewater treatment plants operators and **IGUDAN** (http://www.igudan.org.il) for their help in the sampling efforts.

